# Use of a Self-Care and Educational Mobile App to Improve Outcomes of Patients with Acute Decompensated Heart Failure during the COVID-19 Pandemic

**DOI:** 10.1101/2022.07.28.22270513

**Authors:** Hani Essa, Carolyn Jackson, Siji Nyjo, Ann-Marie Kelly, Naomi Murphy, Nick Hartshorne-Evans, Lauren Walker, Emeka Oguguo, Homeyra Douglas, Rajiv Sankaranarayanan

**Affiliations:** Aintree University Hospital, Liverpool Centre for Cardiovascular Science, Liverpool University Hospitals NHS Foundation Trust, Lower Lane Liverpool L9 7AL United Kingdom; Aintree University Hospital, Liverpool University Hospitals NHS Foundation Trust, Lower Lane Liverpool L9 7AL United Kingdom; The Pumping Marvellous Foundation Heart Failure Patient Charity, Suite 111 Business First Millenium City Park, Millennium Road, Preston PR2 5BL United Kingdom; University of Liverpool, Liverpool University Hospitals NHS Foundation Trust, Prescot St, Liverpool L7 8XP United Kingdom; Liverpool University Hospitals NHS Foundation Trust, Lower Lane Liverpool L9 7AL United Kingdom; NIHR CRN Research Scholar and Honorary Senior Clinical Lecturer (University of Liverpool), Liverpool Centre for Cardiovascular Science, Liverpool University Hospitals NHS Foundation Trust, Lower Lane Liverpool L9 7AL United Kingdom

**Keywords:** Heart failure, App, Self-care, medication, hospitalisation

## Abstract

**Introduction:** Expansion in digital health using mobile phone health applications has increased recently. We developed a mobile phone application (Aintree Heart Failure Passport-AHFP APP) for heart failure (HF) patient education, self-care and improved medication adherence.

**Methods:** This was a prospective observational study of patients with acute decompensated HF managed with day-case intravenous diuretics in a HF specialist nurse delivered Ambulatory Acute Heart Failure Unit (AAHFU) in a British university hospital during the ongoing COVID-19 pandemic (March 2020 to July 2021). We assessed self-care behaviour (European Heart Failure Self-care Behaviour scale – EHFSBs-9) and medication adherence (Medication Adherence Report Scale -MARS-5) at 2 weeks post-presentation in patients who utilised the AHFP APP and compared 30-day HF re-admissions with annual hospital HF data.

**Results:** 148 out of 221 consecutive ADHF patients treated in the AAHFU downloaded the AHFP Mobile APP. 45% were women and mean age of the cohort 62 ± 6.1 years. 55% patients had HF with reduced ejection fraction (HFrEF), 34% had HF with preserved EF (HFpEF) and 11% had HF with mildly reduced EF. Mean EHFSBs-9 was 19.1±6.7; mean MARS-5 score 23.3±1.HF 30 day re-hospitalisation incidence significantly lower (11%) in the APP cohort compared to the incidence of 19% amongst all patients with ADHF during the study period (p=0.02).

**Conclusions:** Our pilot feasibility study suggests that use of a HF educational self-care mobile phone APP in ADHF patients during the COVID pandemic, leads to high quality self-care behaviour, high medication adherence and also lower levels of 30-day HF re-hospitalisation. These results will need to be validated in a randomised controlled trial.

**3 Key Points:**

- The use of digital healthcare technologies such as mobile APPs, is rapidly increasing
- This study analyses the role of our heart failure educational and self-care mobile APP, used by patients with acute decompensated heart failure during the COVID-19 pandemic
- Our results show that use of the mobile APP can lead to high levels of self-care, medication adherence and also reduced 30 day readmissions

## Introduction

Heart failure (HF) is a global cause of significant morbidity as well as mortality, with an estimated prevalence of 1 million patients in the UK and 26 million patients worldwide^1^. Patients with HF are at a high risk of readmission following index hospitalisation (25% within one month) and hospitalisations consume the bulk of costs (nearly 75%) related to HF care^2^. Overall, the management of HF is estimated to cost around $30.7 billion in 2012 in the US and this is expected to balloon to $69.7 billion by 2030^1^. Therefore, there is a pressing need for novel strategies aimed at improving HF care in a cost-efficient manner.

More than 50% of emergency hospital admissions for HF are associated with moderate or severe oedema^3^. As peripheral oedema usually accumulates over days or weeks and leads to weight gain, there is a clear opportunity to reduce emergency hospital admissions through patient self-management, better identification and control of fluid overload in the community.

We developed the Aintree Heart Failure Passport Mobile Application (App) for HF patient self-education and improved medication adherence. The aim of this study is to assess the effect of our mobile HF app upon self-care behaviour, medication adherence and 30 day re-hospitalisation during the ongoing Coronavirus disease-19 (COVID-19) pandemic.

## Methods

This study was conducted in British university hospital from March 2020 to July 2021 during the ongoing COVID-19 pandemic. We assessed self-care behaviour (European Self-Care Behaviour Scale-9^4^) and medication adherence (MARS-5)^5^ at 2 weeks following attendance with ADHF amongst patients in the Ambulatory Acute HF Unit (AAHFU) who utilised a dedicated HF mobile Application (APP) (Aintree Heart Failure Passport Mobile APP). The AAHFU is a HF nurse delivered, consultant-led multidisciplinary day-case unit for out-patient management of ADHF with bolus intravenous diuretics.

The AHFP Mobile APP is available on both iOS (https://apps.apple.com/gb/app/aintree-heart-failure-passport/id1410856426) as well as Android platforms (https://play.google.com/store/apps/details?id=com.s3kdevelopers.aintreeheartfailurepassport&hl=en_GB&gl=US), free of charge. The APP incorporates the following patient education and self-care strategies -

- Traffic Light Based Tracker of patient symptoms and self-care advice (Figure)
- Daily weight monitoring and trigger for contacting HF team based on increased weight due to fluid overload
- Improved medication adherence through timed alerts/prompts as well as alerts about healthcare appointments
- Improved patient education regarding HF using videos
- Mood-check questionnaire to screen for anxiety or depression

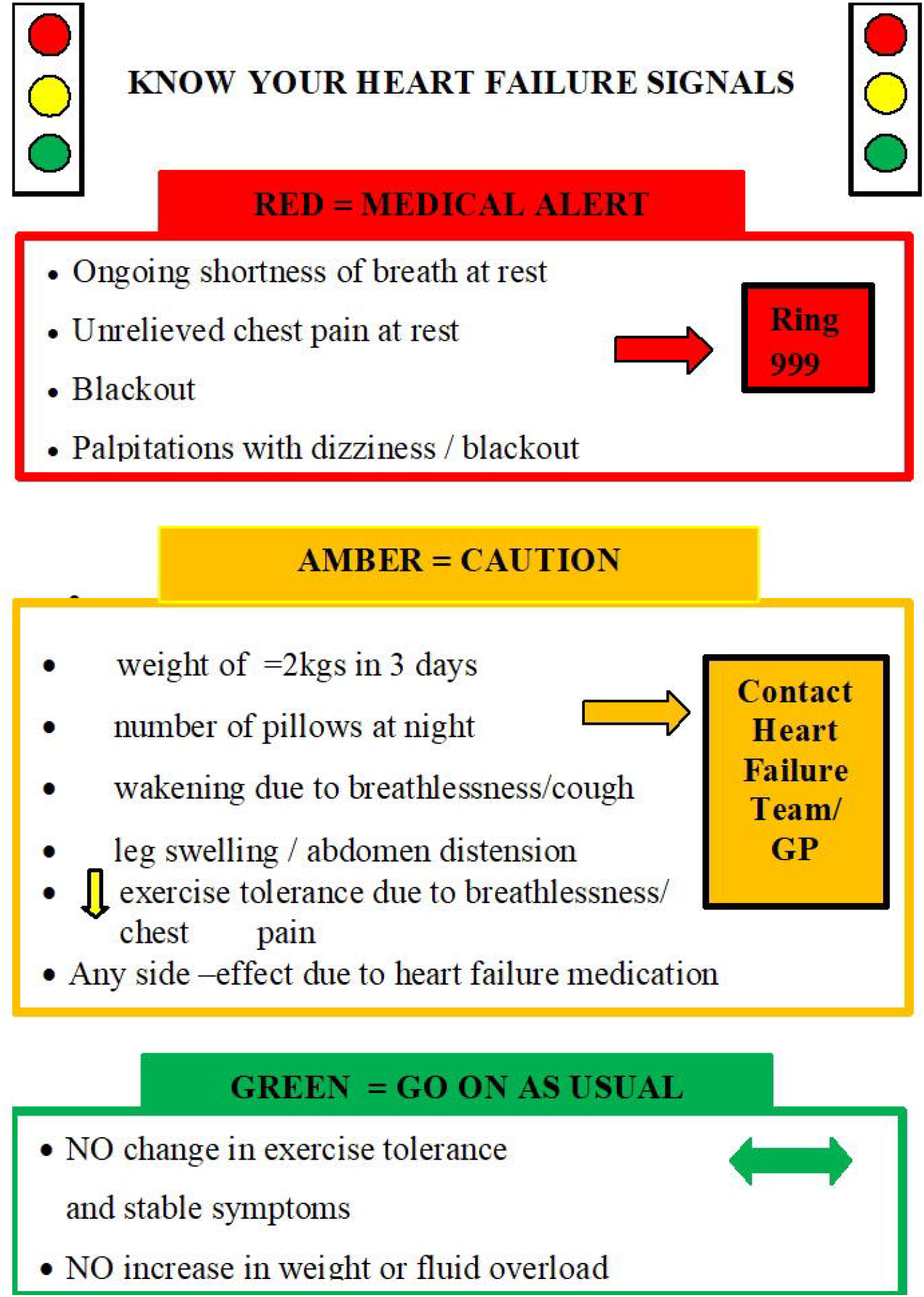

Our study included patients from March 2020 to July 2021 who presented to the AAHFU with a primary diagnosis of acute decompensated heart failure (ADHF) based on the European Society of Cardiology definition of acute heart failure^6^. Other inclusion criteria included an ability to speak, understand and read English, possess a smartphone or have a carer who was agreeable to use the mobile App on their smartphone. Exclusion criteria consisted of cognitive impairment or those who died during the index admission.

Descriptive statistics were represented as continuous variables as means with standard deviations. Categorical variables were represented with percentages and analysed using Chi-squared test. Our study complied with the Declaration of Helsinki; the mobile App (funded by the hospital) and protocol were approved by the Institutional Board. The HF App study group included HF consultant cardiologists, HF specialist nurses, cardiology fellows, cardiac pharmacist, consultant pharmacologist and a HF expert patient.

## Results

Outcomes of 148 out of 221 (67%) consecutive ADHF patients were analysed. 6% were excluded due to cognitive impairment, 20% did not have access to a smart phone and 7% were excluded due to being on the palliative care pathway. 45% of the recruited patients were women and the mean age of the cohort was 62 ± 6.1 years. 55% patients had HF with reduced ejection fraction (HFrEF), 34% had HF with preserved EF (HFpEF) and 11% had HF with mildly reduced EF (HFmrEF). The Charlson Co-morbidity Index^7^ in this cohort was moderate to severe (4.9 ± 1) and frailty (as assessed by the Rockwood Clinical Frailty Scale^8^ was mild (mean 4.7 ± 0.7).

HF self-care behaviour as assessed by the mean EHFSBs-9 was 19.1±6.7 indicating a high level of self-care behaviour. Similarly medication adherence as assessed by the mean MARS-5 score was also good (mean score 23.3±1). In total 16/148 (11%) patients were re-hospitalised within 30 days with HF (73 bed-days) compared to the annual hospital HF rehospitalisation rate which was 19%.

## Discussion

Our study demonstrates that use of our HF mobile App can lead to improvements in several HF outcomes, especially high levels of self-care and high levels of medication adherence. It is likely that these in turn led to lower rates of 30-day HF re-hospitalisation. Whilst previous studies have shown the importance of nurse-led HF self-education on improved outcomes^9^, this is the first study reporting the results of HF patient education through use of a mobile phone APP, during the COVID-19 pandemic. These results are in keeping with a large review of 18 studies looking at a variety of HF Apps demonstrating trends towards a positive impact in patient care^10^.

A 2017 position statement from the American Health Association (AHA) advised that that self-care research has been held back by the perception by both physicians and patients that pharmacological interventions are more effective than lifestyle change despite evidence to the contrary and moving forward, a greater emphasis should be placed on self-care in evidence-based guidelines^11^. The use of emerging health technologies such as smart devices to deliver health care services has been consistently on the rise over the past decade^12^. The pervasive and ubiquitous nature of smart phones and the Apps that define them, create an opportunity in healthcare for closer integration of physicians and their patients. Smartphone Apps can create innovative solutions such as remote education and promotion of self-care strategies to help improve the outcomes of long term conditions such as HF. This is particularly relevant in the current era of the COVID-19 pandemic where mobile healthcare education can help reduce hospital visits and reduce the risk of further disease spread.

## Conclusions

There is currently a rapidly growing requirement for digital technology interventions that can help promote heart failure self-care and thereby improve patient outcomes. Results from our pilot study suggest that use of a HF self-care mobile phone app (Aintree Heart Failure Passport App) in patients with ADHF, leads to high quality self-care behaviour, high levels of medication adherence and lower levels of 30-day HF re-hospitalisation. These results will need to be further validated in a randomised controlled trial.

## Data Availability

All data produced in the present study are available upon reasonable request to the authors

## Acknowledgement of grant support

The Aintree Heart Failure Passport was developed through grant of £8000 awarded to Dr Sankaranarayanan by the Directors’ Dragon’s Den from Aintree University Hospital

